# Subtypes of Internalizing and Externalizing Problems in Autistic Preschool Children: Participation in Daily Life and Family Outcomes

**DOI:** 10.64898/2026.04.14.26350723

**Authors:** Takuto Nakamura, Iku Koshio, Hirofumi Nagayama

**Author notes:** **Correspondence**: Takuto Nakamura, Department of Occupational Therapy, School of Health and Welfare Sciences, Kanagawa University of Human Services, 1-10-1 Heisei-cho, Yokosuka-shi, Kanagawa 238-8522, JAPAN.

## Abstract

**Aim:** Autistic children have a high but varied prevalence of internalizing and externalizing problems. This study aimed to identify the subtypes of internalizing and externalizing problems among autistic preschool children in Japan, examine their temporal stability, and investigate differences in participation in daily life and family outcomes across these subtypes.

**Methods:** A prospective cohort study was conducted with 275 caregivers of autistic children aged 51–75 months. Internalizing and externalizing problems were assessed using the Strengths and Difficulties Questionnaire.

**Results:** Latent transition analysis identified five subtypes: Low-symptom, High-emotional, Externalizing, Comorbid, and Peer-difficulty groups. Membership in the High-emotional and Externalizing groups was relatively stable over time, whereas the Peer-difficulty group showed frequent transitions to subtypes with higher levels of internalizing or externalizing problems. Significant differences in participation in daily life and family outcomes were observed across subtypes, but these patterns were inconsistent with a simple gradient of symptom levels.

**Conclusions:** The novel findings that the temporal stability of subtype membership varied and that differences in participation in daily life and family outcomes were observed across the subtypes suggest that the heterogeneity of internalizing and externalizing problems may be associated with variations in children’s participation in daily life and family outcomes over time.

**Plain Language Summary:** Autistic preschool children often experience emotional and behavioral difficulties, but the way these difficulties manifest varies widely across individuals. This study aimed to identify the patterns of these difficulties, examine how they change over time, and investigate how participation in daily life and family outcomes differ across autistic preschool children. We conducted a study with 275 caregivers of autistic children aged 4–6 years in Japan. From caregiver reports of children’s emotional and behavioral difficulties, five distinct patterns were identified: a group with mainly emotional difficulties, a group with mainly behavioral difficulties, a group with both types of difficulties, a group with relatively low levels of difficulties, and a group characterized primarily by peer-related difficulties. Our findings suggest that different patterns of emotional and behavioral difficulties are associated with differences in children’s participation in daily life and family outcomes. These differences could not be explained simply by the overall severity of difficulties but rather reflect distinct patterns based on the type of difficulty. The results indicate that autistic children face diverse difficulties that change over time.

## Introduction

Autistic children exhibit higher rates of internalizing and externalizing problems than those without autism (Salazar et al., 2015). Internalizing problems refer to inwardly directed difficulties such as anxiety, depression, and social withdrawal, whereas externalizing problems refer to outwardly directed behavioral issues such as impulsivity, aggression, and hyperactivity (Goodman et al., 2010). Mental health problems in childhood that include internalizing and externalizing difficulties are associated with developmental impairments such as poor academic performance (Lee et al., 2007), and internalizing and externalizing problems during the developmental period have been shown to predict an increased risk of work incapacity in young adulthood (Narusyte et al., 2017). A variety of factors are implicated in the emergence of internalizing and externalizing problems, including language ability (Levy et al., 2023), sensory processing characteristics (Chen et al., 2024), and parental mental health (Yorke et al., 2018).

Traditional analytic approaches have limitations in addressing the wide heterogeneity of internalizing and externalizing problems among children. Some children show co-occurring internalizing and externalizing problems, whereas others exhibit only one of these difficulties or show minimal problems overall (Vaillancourt et al., 2017). As this diversity is difficult to capture through simple group comparisons, person-centered statistical methods such as latent class analysis and latent class growth curve modeling are increasingly used for subtyping (Nylund-Gibson et al., 2023; Petersen et al., 2019). Subtyping offers substantial clinical value because it allows for more precise examinations of associated and risk factors, enabling individualized support planning and risk assessment rather than treating internalizing and externalizing problems as uniform.

Subtyping studies of internalizing and externalizing problems have primarily used caregiver-reported questionnaires to collect data. For example, Piergies et al. (2022) examined subtypes of internalizing and externalizing problems among autistic schoolchildren using latent class analysis and identified four groups: a Mild Psychiatric Symptoms group, a group characterized by prominent internalizing problems, a group characterized by prominent externalizing problems, and a High Psychiatric Symptoms group in which both internalizing and externalizing problems were markedly elevated. They also demonstrated that these four subtypes differ in terms of their IQ, sleep problems, and prevalence of co-occurring medical conditions. Vaillancourt et al. (2017) conducted subtyping based on the joint developmental trajectories of internalizing and externalizing problems in autistic preschool children using latent class growth analysis, which allowed them to account for changes over time. Approximately 40% of children were classified into a low-risk group that exhibited persistently low levels of both internalizing and externalizing problems, whereas 13–15% were classified into a high-risk group in which both problem types remained consistently elevated. The remaining children demonstrated developmental trajectories in which either internalizing or externalizing problems were relatively more pronounced, yielding a distribution pattern that largely aligns with the single time point subtype profiles of Piergies et al. (2022). Taken together, these studies suggest that subtyping can be examined in terms of not only symptom profiles at a given time point but also whether such patterns sustain over time.

However, research in this area remains limited for two main reasons. First, most subtyping studies of internalizing and externalizing problems have been conducted primarily in the United States and Canada and whether these findings generalize to regions with different cultural backgrounds is unclear. As the factors associated with internalizing and externalizing problems are likely to be closely tied to sociocultural contexts such as support systems, educational environments, and family culture, examining autistic children in a country with different social systems and educational settings could provide valuable insights into the generalizability of subtype patterns and associated factors (Nakamura, Dobashi, et al., 2025).

Second, the associated factors examined in previous subtyping studies have largely been limited to individual characteristics such as sex, IQ, and core autism symptoms, with little attention paid to contextual aspects such as living environments and family-related variables (Piergies et al., 2022; Vaillancourt et al., 2017). For example, participation in daily life is a central concept in supporting autistic children and their families, and clarifying the relationship between internalizing and externalizing problems and enhanced participation in everyday activities could yield important knowledge for developing individualized support plans (Danielsson et al., 2024). Furthermore, family support is a key focus of early intervention for autistic children (Yorke et al., 2018). Family outcomes serve as indicators that capture the effects of support services on caregivers’ psychological well-being, parenting responsibility, and daily functioning, making them valuable for understanding families’ overall situations (Ueda et al., 2015). However, few studies have examined the associations between family outcomes and internalizing and externalizing problems across subtypes. Moreover, studies have not directly examined the stability of subtype membership over time. Therefore, it remains unclear whether the subtype patterns identified in prior studies represent relatively stable behavioral profiles or whether children transition between subtypes as they develop.

Based on these gaps in the literature, the present study aimed to identify subtypes of internalizing and externalizing problems among autistic children in Japan—where social systems and educational settings differ from those in North America—to examine the temporal stability of these latent subtypes over time as well as investigate differences across subtypes in children’s participation in daily life and family outcomes.

## Methods

This study was a prospective cohort study conducted across four time points (Times 1–4) between July 2023 and February 2024. Times 1 and 2 were conducted between July and August 2023 with an interval of approximately one week, Time 3 was conducted approximately three months later, and Time 4 was conducted approximately three months after Time 3.

### Participants

The inclusion criteria were as follows: (1) being a caregiver of a preschool-aged child between 51 and 75 months, (2) having a child diagnosed with Autism spectrum disorder (ASD) by a physician, and (3) having a child with an IQ or developmental quotient of 50 or higher based on recent caregiver-reported scores from eligible tests, including the Wechsler Intelligence Scale for Children (WISC), Wechsler Preschool and Primary Scale of Intelligence (WPPSI; Wahlstrom et al., 2018), Tanaka–Binet Intelligence Scale (TBIS; Uno et al., 2014), and Kyoto Scale of Psychological Development (KSPD; Koyama et al., 2009). The exclusion criterion was having a child diagnosed by a physician with a condition other than a neurodevelopmental disorder; children with comorbid neurodevelopmental disorders were included, whereas those with comorbid non-neurodevelopmental disorders were excluded.

### Measures

#### Strengths and Difficulties Questionnaire (SDQ)

The SDQ is an instrument for screening children’s mental health that assesses five subdomains: emotional symptoms, conduct problems, hyperactivity/inattention, peer problems, and prosocial behavior. The Japanese parent-report version of the SDQ for ages 4–16 was used in the present study (Matsuishi et al., 2008). The internal consistency of the SDQ is acceptable for the total difficulties score (Cronbach’s α = 0.77). By subdomain, internal consistency was acceptable for hyperactivity/inattention (α = 0.75), questionable for emotional symptoms (α = 0.61) and prosocial behavior (α = 0.69), and poor for conduct problems (α = 0.52) and peer problems (α = 0.52). Higher scores indicate greater difficulties on all the subdomains except prosocial behavior, for which higher scores indicate better functioning.

#### Participation Questionnaire for Preschoolers (PQP)

The PQP assesses the participation in daily life of autistic children over the past three months across four subdomains: friendships and education, family satisfaction, daily life and independence, and leisure and community life (Nakamura, Nagayama, et al., 2025). The PQP has acceptable internal consistency (Cronbach’s α = 0.74–0.88 for the subdomains; α = 0.83 for the total score), test–retest reliability (intraclass correlation coefficient = 0.84–0.92 for the subdomains; intraclass correlation coefficient = 0.93 for the total score), structural validity (verified through exploratory factor analysis), construct validity, and responsiveness (with 75% of hypotheses supported) (Nakamura et al., 2026; Nakamura, Nagayama, et al., 2025). Higher scores indicate greater participation. Because the present study was conducted concurrently with the PQP scale development study, a 36-item trial version (from which 7 items were later removed) was used (Nakamura et al., 2024), although the deleted items were not included in the final scoring.

#### Japanese Version of the Social Responsiveness Scale, Second Edition (SRS-2-J)

The SRS-2 evaluates the severity of autismsymptoms (Constantino, 2017). The child version used in this study (for ages 4–18) is a caregiver-report measure comprising five subdomains: social awareness, social cognition, social communication, social motivation, and restricted interests and repetitive behavior. The total score, calculated by summing all five subdomain scores, is converted into sex-standardized T-scores. The psychometric properties of the SRS-2-J have not yet been systematically validated. However, the English version and multiple other language versions have demonstrated excellent reliability, with Cronbach’s alpha coefficients exceeding 0.90 (Constantino & Gruber, 2017). In the present study, the internal consistency of the SRS-2-J T-scores was excellent (α = 0.93). Higher SRS-2-J scores indicate more severe Autism symptoms.

#### Japanese Version of the Family Outcomes Survey – Revised (FOS-J)

The FOS-J evaluates the effects of support and intervention programs on children and their families (Ueda et al., 2015). The instrument consists of two sections: family outcomes and support efficacy. In the present study, only the family outcomes section was used, which comprises five subdomains: understanding your child’s strengths, needs, and abilities; understanding your rights and advocating for your child; supporting your child’s development and learning; accessing the support system; and accessing community resources. Higher scores indicate that the support services are producing beneficial outcomes for families. The FOS-J has demonstrated structural and construct validity as well as very high internal consistency (Cronbach’s α = 0.94 for the full scale) (Ueda et al., 2015).

#### Check List of Obscure disAbilitieS in Preschoolers (CLASP)

The CLASP screens for stuttering, tic disorders, specific learning disabilities, and developmental coordination disorder (DCD); in the present study, only the five-item DCD subdomain was used (Inagaki, 2017). Response options are “never,” “rarely,” “sometimes,” “often,” and “always.” A child is considered at an increased risk of DCD if any item is endorsed as “often” or “always.” Although the psychometric properties of the CLASP DCD subdomain have not yet been systematically validated, the data provided by participants at baseline in this study indicated acceptable internal consistency (Cronbach’s α = 0.78). When administered to older preschool children, the intraclass correlation coefficients were 0.485 for boys and 0.573 for girls, with caregivers tending to assign lower scores (Inagaki, 2017).

#### Japanese Version of the Kessler Psychological Distress Scale (K6-J)

The K6-J is a tool for screening psychological distress. In the present study, respondents were asked to report their own psychological distress rather than that of their autistic children. The reliability of the K6-J is high (Cronbach’s α ≥ 0.80) (Nishi et al., 2012; Sakurai et al., 2011). The scale consists of six items, with higher total scores indicating poorer mental health (Furukawa et al., 2008).

#### Demographic Information

Demographic information was collected on family composition, educational background, household income, the target child’s sex and age in months, comorbid diagnoses, number of days per week the child used child developmental support services, presence of professional support, the child’s age in months at ASD diagnosis, and the age at which intervention services began. In addition, based on the IQ results from the WISC, WPPSI, and TBIS or the developmental quotient results from the KSPD, cognitive level was classified into intellectual disability (IQ/developmental quotient 50–69), borderline intelligence (70–84), average intelligence (85–114), and high intelligence (115 or above).

### Sample and Data Collection

The sample size was determined based on the goal of obtaining complete responses from at least 300 participants (Nylund-Gibson et al., 2023). Participants were recruited through Hattatsu Navi, a website providing information on developmental disabilities with over 300,000 registered members, as well as through the mailing lists of LITALICO Junior, which operates 147 facilities in Japan, and LITALICO Works, which operates 137 facilities (LITALICO Inc., 2023, 2024). Individuals who expressed an interest were emailed a URL linking to the online survey platform (Questant), where they were invited to complete the questionnaire. Informed consent was obtained through the platform.

The survey was conducted over four time points (Times 1–4) between July 2023 and February 2024 to avoid the institutional transition period associated with the start of the academic year in April in Japan. As some participants may be registered on multiple mailing lists, the informed consent process explicitly stated that duplicate participation was prohibited. After data collection was completed, the first and second authors confirmed the absence of duplicate entries by checking children’s birthdates, regions of residence, and dates of ASD diagnosis. Additionally, responses to the PQP were reviewed at all time points, and the two authors confirmed that no participants had provided uniform responses across all items (e.g., selecting “1” for every item).

### Data Analysis

Using all the SDQ subdomains except prosocial behavior, Z-scores were calculated based on data from a previous study conducted with non-autistic children in Japan (Matsuishi et al., 2008). Each time point was included in the model as a covariate. One- to seven-class models were tested, and the Bayesian information criterion (BIC) and Akaike’s information criterion (AIC) were used to assess the model fit (Nylund-Gibson et al., 2023). Posterior probabilities indicating the likelihood that each individual belonged to each class were estimated based on the selected number of classes. The class with the highest posterior probability was assigned as the individual’s latent profile.

Subsequently, class differences were examined using χ² tests for the following variables: single-parent household (presence of a spouse), university-level education or higher, household income ≤3 million yen, household income ≥10 million yen, child’s sex, cognitive level, presence of comorbid ADHD, presence of DCD risk according to the CLASP, and use of child developmental support services. For variables showing significant differences, Fisher’s exact test was used for pairwise multiple comparisons, with Bonferroni corrections applied to adjust for Type I error. In addition, Kruskal–Wallis tests were conducted to examine the differences in respondents’ K6-J total scores, child’s age at diagnosis, number of months since ASD diagnosis, and subdomain and total scores for the PQP, SDQ, FOS-J, and SRS-2-J by class. When significant differences were found, Dunn’s multiple comparison tests were performed, with Bonferroni corrections applied to control for Type I error inflation.

To evaluate the transitions between classes at each time point, transition probabilities were estimated using a Markov model. The EM algorithm was used for the estimation, with 500 iterations specified, and computations were performed until parameter convergence was achieved at below 0.01. LatentGOLD 6.0 (Statistical Innovations Inc., Belmont, MA, USA) was used for the latent transition analysis. Between-group comparisons were conducted using R version 4.4.2 (R Core Team, 2024). Dunn’s multiple comparison tests were implemented using the FSA package in R (Ogle et al., 2023).

### Ethical Considerations

The study was approved by the Research Ethics Committee of the [BLINDED FOR REVIEW]. The study was preregistered before data analysis (Nakamura et al., 2024). The preregistration included the study design, survey time points, measurement items, and the main analysis plan.

## Results

### Participants’ Characteristics

A total of 545 caregivers met the criteria and participated at Time 1, of whom 275 completed all four surveys and provided complete data (retention rate: 50.5%). As shown in Table 1, 96% of respondents were mothers. The sample showed higher household income and a higher proportion of individuals with university education than the national average in Japan (Ministry of Health, 2022; Statistics Bureau, 2021). Among the target children, 79% were boys, and autism severity—based on the T-scores of the SRS-2-J—showed a distribution in which most children fell within the moderate to severe range (Table 2) (Constantino, 2017).

**Table 1.**
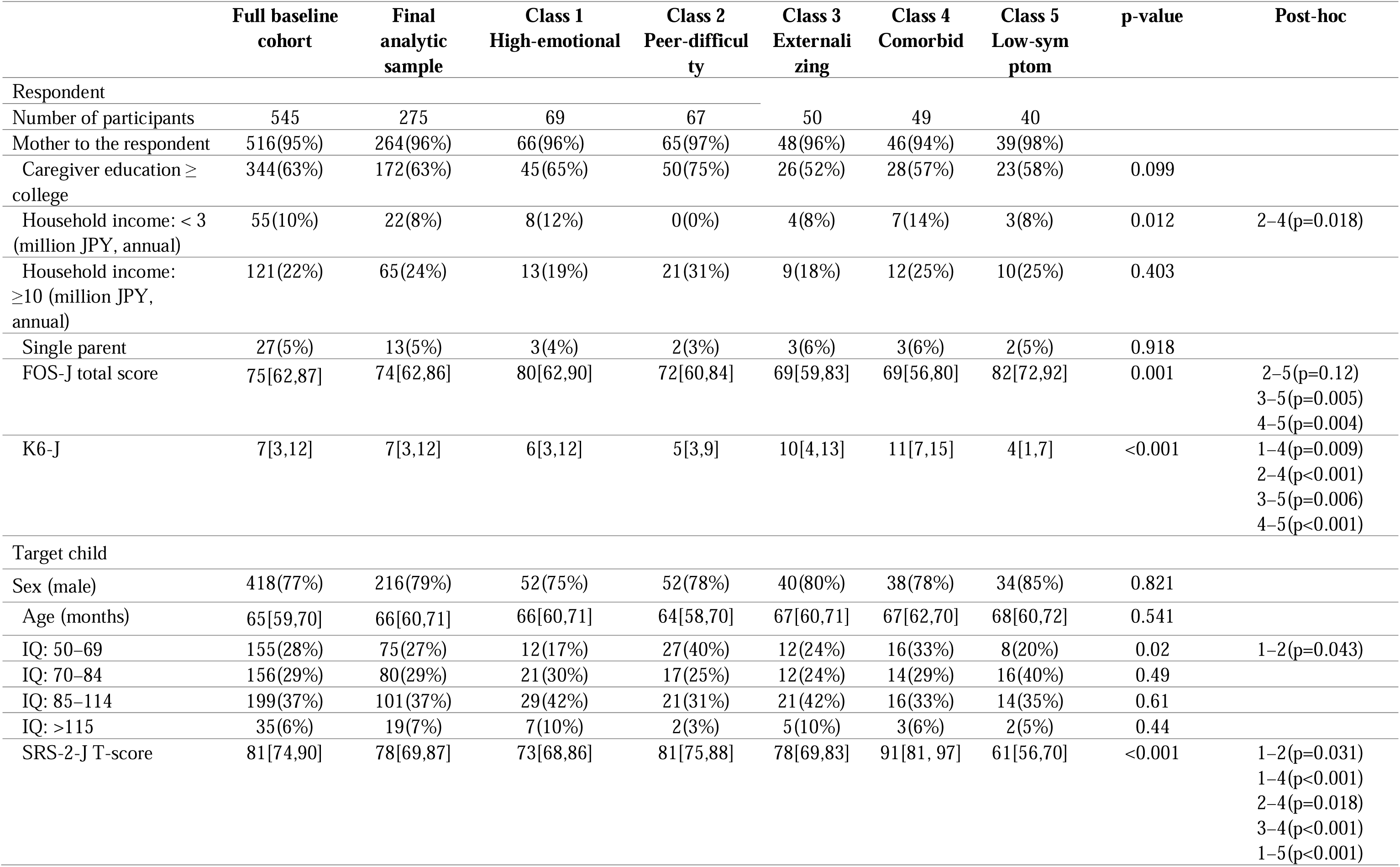
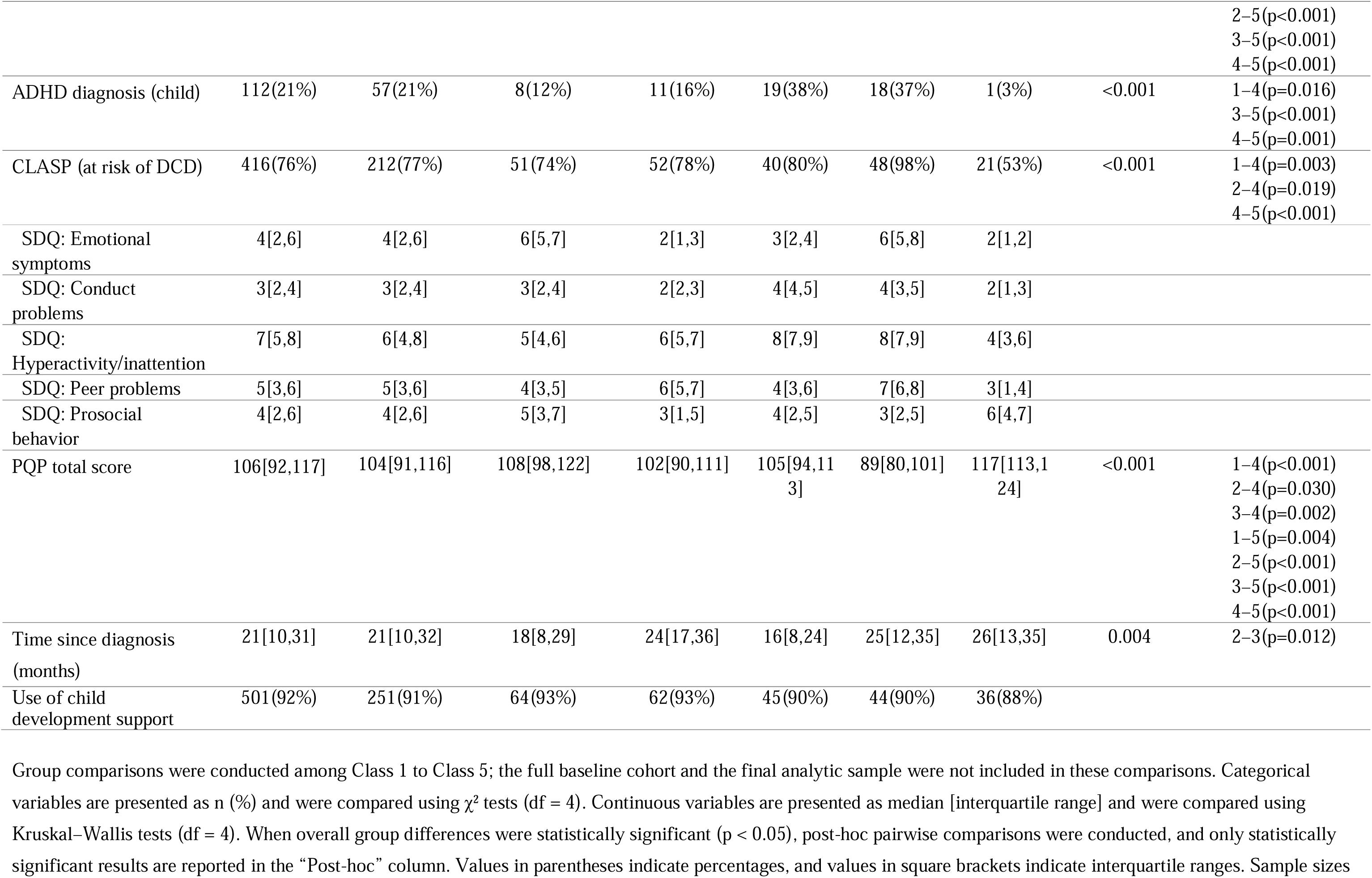
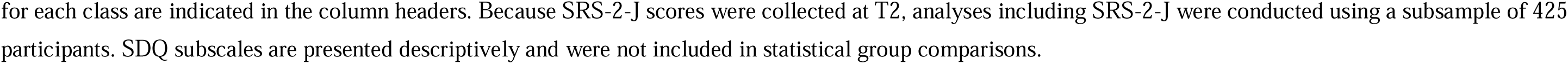
Demographic and clinical characteristics of the participants overall and by class.

**Table 2.**
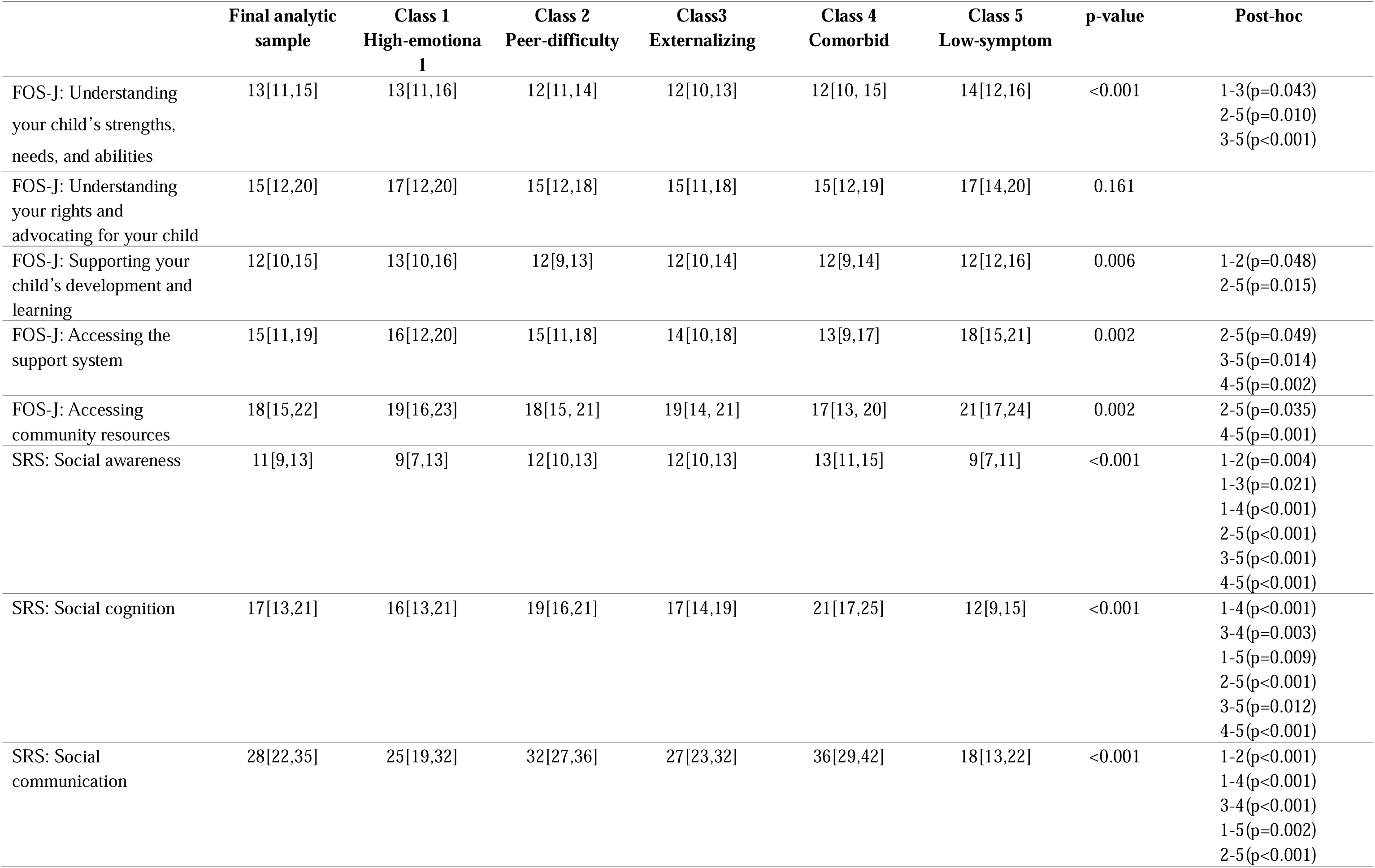
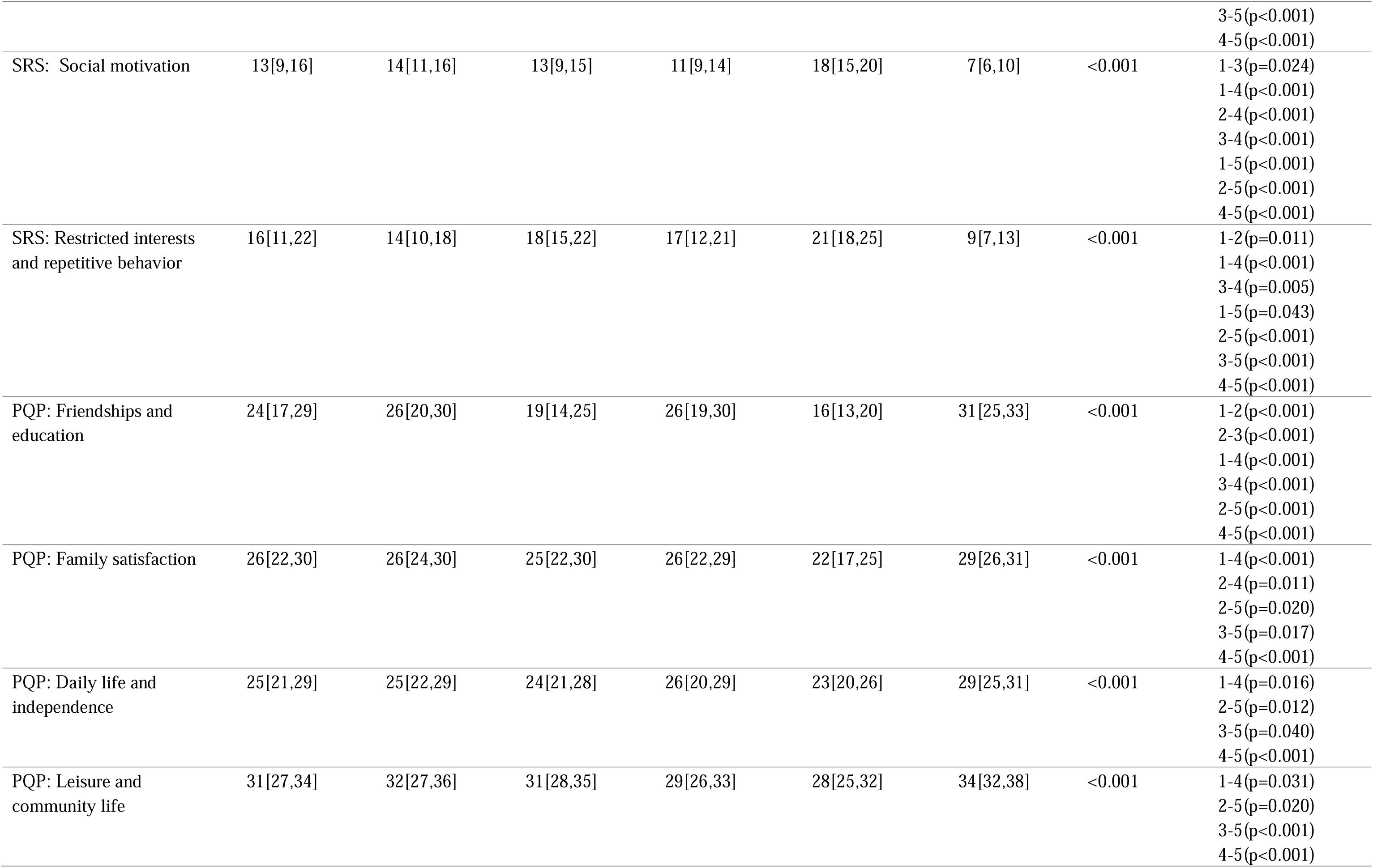
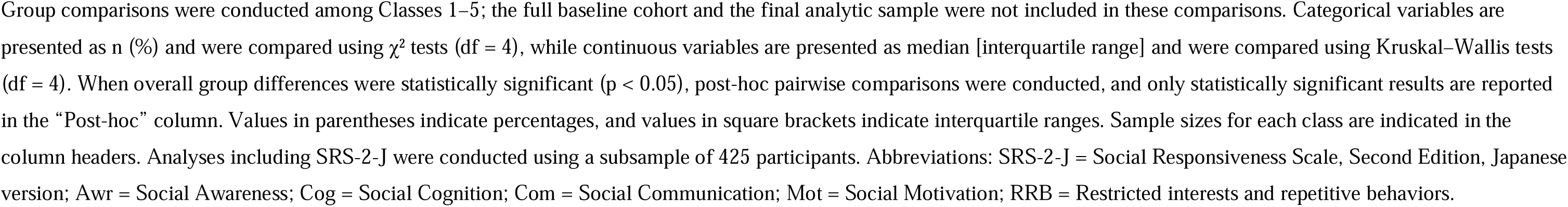
Comparison of the family outcomes, symptom severity, and participation in daily life by class.

The five-class model demonstrated the lowest BIC among the evaluated models and this was adopted as the optimal solution (Table 3). As illustrated in Figure 1, Class 1 showed high emotional symptoms scores and low scores on the externalizing problem domains (conduct problems and hyperactivity/inattention) for the SDQ; it was therefore labeled the High-emotional group. This class tended to include fewer children with intellectual disabilities and had a later age at ASD diagnosis. It showed moderate SRS-2-J T-scores but high total PQP scores and low K6-J scores.

**Figure 1.**
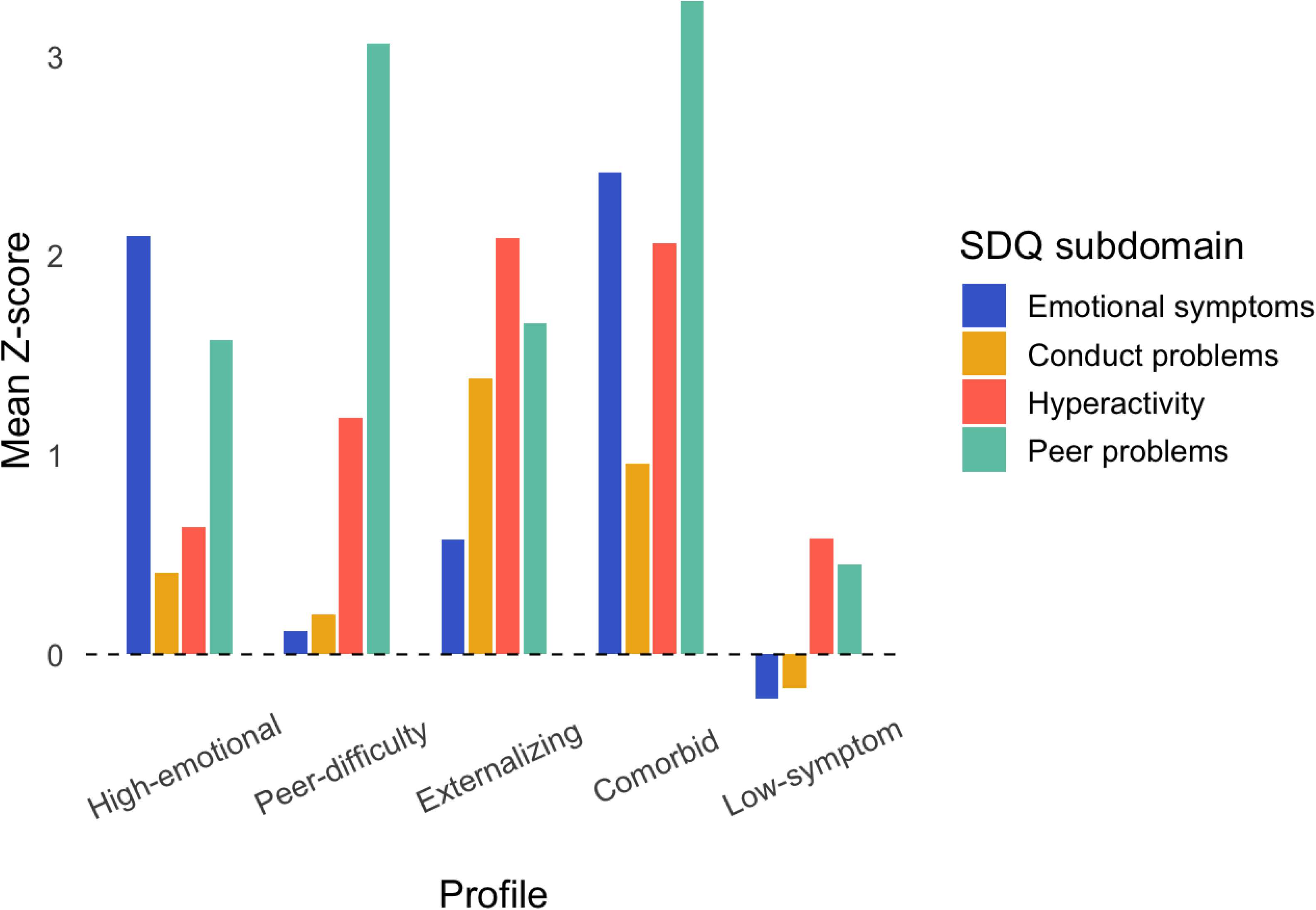
Z-scores of the SDQ subdomains by class.

**Table 3.** Model fit indices for latent profile analysis across one- to seven-class solutions.

| **Class** | **LL** | **BIC** | **AIC** | **Npar** | **Entropy** |
| --- | --- | --- | --- | --- | --- |
| 1-Class | -5283.441 | 10611.816 | 10582.881 | 8 | 1 |
| 2-Class | -5050.230 | 10218.413 | 10142.461 | 21 | 0.862 |
| 3-Class | -4895.361 | 10004.159 | 9866.722 | 38 | 0.843 |
| 4-Class | -4792.398 | 9916.186 | 9702.797 | 59 | 0.873 |
| 5-Class | -4696.832 | 9865.473 | 9561.664 | 84 | 0.877 |
| 6-Class | -4632.522 | 9899.740 | 9491.045 | 113 | 0.886 |
| 7-Class | -4583.430 | 9986.909 | 9458.860 | 146 | 0.892 |

Class 2 demonstrated low to moderate scores on all the subdomains of the SDQ except peer problems and was labeled the Peer-difficulty group (Figure 1). This group tended to include more children with intellectual disabilities and showed relatively high SRS-2-J T-scores. Family outcome scores were generally low, whereas the K6-J scores were low. This subtype was also characterized by an earlier age at ASD diagnosis.

Class 3 showed high scores on externalizing problems and lower scores on internalizing problems in the SDQ; it was therefore labeled the Externalizing group (Figure 1). This subtype exhibited relatively moderate SRS-2-J T-scores but had low total FOS-J scores and high K6-J scores. Further, it showed later ages at ASD diagnosis and a later initiation of medical and welfare services than those in the Peer-difficulty group.

Class 4 showed moderate to high scores on both the internalizing and the externalizing domains in the SDQ and was therefore labeled the Comorbid group (Figure 1). This subtype exhibited high SRS-2-J T-scores, low total PQP and FOS-J scores as well as low scores across the four subdomains of the PQP and the five subdomains of the family outcomes section of the FOS-J, and markedly high K6-J scores. Although the timing of ASD diagnosis was average, the initiation of medical and welfare services occurred earlier.

Class 5 showed the lowest scores for both internalizing and externalizing problems and was therefore labeled the Low-symptom group (Figure 1). This subtype demonstrated low SRS-2-J T-scores, high PQP and FOS-J total scores as well as high scores across the four subdomains of the PQP and the five subdomains of the family outcomes section of the FOS-J, and low K6-J scores.

Overall, significant differences by class were observed for all the subdomains of the SRS-2-J, PQP, and FOS-J, except for the understanding your rights and advocating for your child subdomain of the FOS-J (Table 2).

Table 4 presents the estimated transition probabilities by class at Times 3 and 4, and Appendix 1 shows the major observed transition patterns by class membership. The High-emotional and Externalizing groups showed relatively stable subtype membership, whereas the other groups exhibited transitions into different subtypes over time. In particular, the Peer-difficulty group showed a relatively high frequency of transitions toward subtypes characterized by increased internalizing or externalizing problems. Although the Peer-difficulty group initially showed relatively low levels of difficulties outside the peer problems subdomain, the results suggest that other problems may increase or become more apparent over time.

**Table 4.** Transition probabilities between the latent classes.

| Time t | From class<br>(t-1) | To class (t) |  |  |  |  |
| --- | --- | --- | --- | --- | --- | --- |
|  |  | 1 | 2 | 3 | 4 | 5 |
| 3 | 1 | 0.998 | 0.001 | 0.001 | 0.001 | 0.001 |
| 3 | 2 | 0.000 | 0.926 | 0.072 | 0.001 | 0.001 |
| 3 | 3 | 0.002 | 0.001 | 0.983 | 0.001 | 0.015 |
| 3 | 4 | 0.001 | 0.001 | 0.058 | 0.940 | 0.000 |
| 3 | 5 | 0.001 | 0.001 | 0.001 | 0.001 | 0.998 |
| 4 | 1 | 0.989 | 0.006 | 0.001 | 0.001 | 0.004 |
| 4 | 2 | 0.023 | 0.920 | 0.001 | 0.053 | 0.004 |
| 4 | 3 | 0.000 | 0.001 | 0.998 | 0.001 | 0.001 |
| 4 | 4 | 0.076 | 0.001 | 0.002 | 0.920 | 0.001 |
| 4 | 5 | 0.053 | 0.001 | 0.004 | 0.037 | 0.905 |

Values represent the estimated transition probabilities between the latent states. Rows indicate the latent class membership at time t-1 and columns indicate the latent class membership at time t. Probabilities in each row sum to 1.

## Discussion

This study identified five subtypes of internalizing and externalizing problems among autistic preschool children in Japan and examined how these subtypes differed across a range of background factors, including child characteristics, family outcomes, and participation in daily life.

Subtypes of Internalizing and Externalizing Problems and Temporal Stability

The presented findings are broadly consistent with those of previous research. Among the five subtypes identified in this study, the Low-symptom group, High-emotional group, Externalizing group, and Comorbid group generally corresponded to the Low Psychiatric Symptoms, Internalizing Symptoms, Externalizing Symptoms, and High Psychiatric Symptoms subtypes identified in autistic school-aged children (Piergies et al., 2022). Moreover, similar patterns were reported in a longitudinal study on autistic preschool children focusing on initial levels of internalizing and externalizing problems (Vaillancourt et al., 2017).

By contrast, the Peer-difficulty group represents a distinctive subtype that has not been reported as an independent class in previous research, perhaps because of differences in the measurement tools used. Previous studies have used the Child Behavior Checklist (Piergies et al., 2022; Vaillancourt et al., 2017), whereas the present study employed the SDQ. Because the SDQ includes items related to peer relationships, its structure may have led to the extraction of a class characterized by difficulties specifically in social relationships. Moreover, considering that these peer-related items conceptually overlap with the core social difficulties of autism and that the Peer-difficulty group included a relatively high proportion of children with a lower IQ or developmental quotient, this group may represent a subtype in which psychiatric symptoms and developmental characteristics are inseparably reflected (Ivarsson et al., 2025). Indeed, when the proportions of the Peer-difficulty and Low-symptom groups—both of which exhibit relatively mild internalizing and externalizing problems—are combined, the subtype distribution in this study closely approximates the proportion of mild internalizing and externalizing symptom groups reported by those previous studies (Piergies et al., 2022; Vaillancourt et al., 2017).

From the perspective of temporal stability, the findings suggest that the Peer-difficulty group does not represent a stable subtype characterized by consistently low levels of internalizing and externalizing problems. Rather, its members face peer relationship difficulties that may differentiate into internalizing or externalizing problems as development progresses. In other words, this group may represent a stage preceding the manifestation of internalizing and externalizing problems or a risk state for their emergence. This finding suggests that the subtypes of internalizing and externalizing problems observed in early childhood are not necessarily fixed and may change over the course of development in some children. Therefore, even when children do not initially exhibit pronounced internalizing and externalizing problems, some such as those in the Peer-difficulty group may constitute a developmentally plastic subgroup at an increased likelihood of transitioning toward more severe symptom profiles over time.

### Differences in Demographic Characteristics by Subtype

Although the analytical approaches and target age groups differed, the findings align with those of previous research in that children with an IQ below 70 were more frequently included in the Peer-difficulty group, which exhibited relatively low levels of internalizing and externalizing problems (Piergies et al., 2022). However, this observation may also reflect that caregiver-rated scales underestimate symptoms due to the difficulties faced by children with a lower IQ verbalizing internal states or expressing emotions (Ivarsson et al., 2025). With respect to comorbid neurodevelopmental disorders other than ASD, differences were observed across the subtypes, with those children exhibiting more severe autism symptoms tending to show higher levels of internalizing and externalizing problems. At the same time, the scores of the SRS-2-J subdomains revealed distinct symptom patterns that could not be explained by a simple severity gradient. Furthermore, ADHD diagnoses were more common in the Externalizing and Comorbid groups, while DCD risk was more frequent in the Comorbid group. These findings suggest that although internalizing and externalizing problems partially overlap with neurodevelopmental characteristics, they are not only indicators of overall severity but also represent heterogeneous features that differentiate the subtypes.

Although studies have examined the associations between certain family outcomes (e.g., parenting self-efficacy) and internalizing and externalizing problems (Yorke et al., 2018), researchers have not thus far investigated these associations separately for each subtype of internalizing and externalizing problems among autistic children. In the present study, the SDQ-based subtypes generally showed a pattern in which the greater severity of internalizing and externalizing problems was associated with poorer caregiver psychological well-being. In particular, the Comorbid group exhibited markedly high K6 scores, consistent with earlier findings that greater internalizing and externalizing symptom severity among children is associated with increased caregiver responsibility (Yorke et al., 2018). While this result supports the existence of an association between the severity of children’s internalizing and externalizing problems and caregivers’ psychological distress, it does not rule out that caregiver distress may influence caregiver-reported evaluations of children’s mental health (Ivarsson et al., 2025). By contrast, the distribution of FOS-J scores, which represent family outcomes, displayed a pattern distinct from that of the K6. For example, in the Peer-difficulty group, children’s internalizing and externalizing problems were relatively mild and caregivers’ psychological distress was low, yet family outcomes were poor. This indicates that even when caregiver psychological distress is low, families may still experience challenges in managing daily life or engaging with support systems. Taken together, these findings suggest that the subtypes of children’s internalizing and externalizing problems do not merely reflect the degree of family difficulty; rather, the patterns linking caregiver psychological distress and family outcomes may differ across the subtypes.

Few studies have systematically examined how patterns of internalizing and externalizing problems relate to differences in participation in daily life (Danielsson et al., 2024). The present study adds important value in that it investigates the association between SDQ-based internalizing and externalizing subtypes and participation in daily life separately by subtype and by the subdomains of participation in daily life. The Low-symptom group consistently showed the highest participation across all the subdomains, whereas the Comorbid group showed the lowest levels. Additionally, the severity of internalizing and externalizing problems did not show a linear relationship with reduced participation in daily life. Specifically, in the Externalizing group, participation in the subdomain of friendships and education was relatively preserved despite the presence of externalizing problems, indicating that externalizing symptoms do not necessarily reduce participation in these areas. By contrast, the Peer-difficulty group showed selectively reduced participation in those subdomains related to peer relationships, reflecting a correspondence between subtype characteristics and the specific areas in which participation restrictions emerged. These findings suggest that internalizing and externalizing problems do not uniformly restrict participation in daily life based solely on symptom severity; rather, the subdomains in which participation is more likely to decline differ across the subtypes. Although studies have reported associations between internalizing problems and reduced participation in daily activities (Ambrose et al., 2022), they have overlooked the heterogeneity in symptom profiles. The presented findings suggest that the relationship between internalizing and externalizing problems and participation in daily life may differ by subtype, indicating that different combinations of internalizing and externalizing symptoms may influence participation in different subdomains.

### Limitations

First, the SDQ used in this study is standardized for children aged 4 years and older. Some participating children had IQ scores around 50, making it possible that their mental age was below 4 years. Such discrepancies between developmental age and item content may make it difficult for caregivers to recognize certain behavioral characteristics (e.g., peer relationships, rule-following, emotional self-regulation) as deviations because these behaviors may not yet be developmentally expected. Therefore, internalizing and externalizing problems in these children may have been rated lower than their actual levels, potentially introducing underestimation bias. Second, the study targeted a population receiving specific child developmental support services, which requires caution when generalizing the findings to other populations. Third, respondents tended to have higher household incomes and educational backgrounds than the national average in Japan; further investigation is needed before generalizing the findings to groups with more diverse family backgrounds. Finally, the sample size was relatively small, which may have reduced the statistical power.

## Conclusion

This study identified five subtypes of internalizing and externalizing problems among autistic preschool children in Japan. Four of these subtypes—the Internalizing group, Externalizing group, Comorbid group, and Low-symptom group—were largely consistent with profiles reported in previous studies, suggesting that these patterns of internalizing and externalizing problems may emerge consistently from early childhood regardless of the cultural context. By contrast, the temporal stability of these subtypes varied: while the High-emotional and Externalizing groups showed relatively stable trajectories, the Peer-difficulty group demonstrated a tendency to transition toward subtypes with more severe behavioral problems, indicating that subtype membership may change dynamically for some children. This finding suggests that behavioral and emotional heterogeneity in early childhood may change in response to family and environmental factors. The associations among the subtypes, participation in daily life, and family outcomes highlight the need to consider not only child-specific characteristics but also the broader context of daily life when evaluating risk and determining support strategies.

## Data Availability

The data that support the findings of this study are not publicly available due to ethical and privacy restrictions.

## Acknowledgments

We would like to acknowledge the Department of Developmental Disorders, National Institute of Mental Health, National Center of Neurology and Psychiatry (NCNP), for granting permission to use the CLASP (Check List of Obscure disAbilitieS in Preschoolers).

We also thank Editage (www.editage.com) for English language editing and submission support.

---

**Appendix 1.**
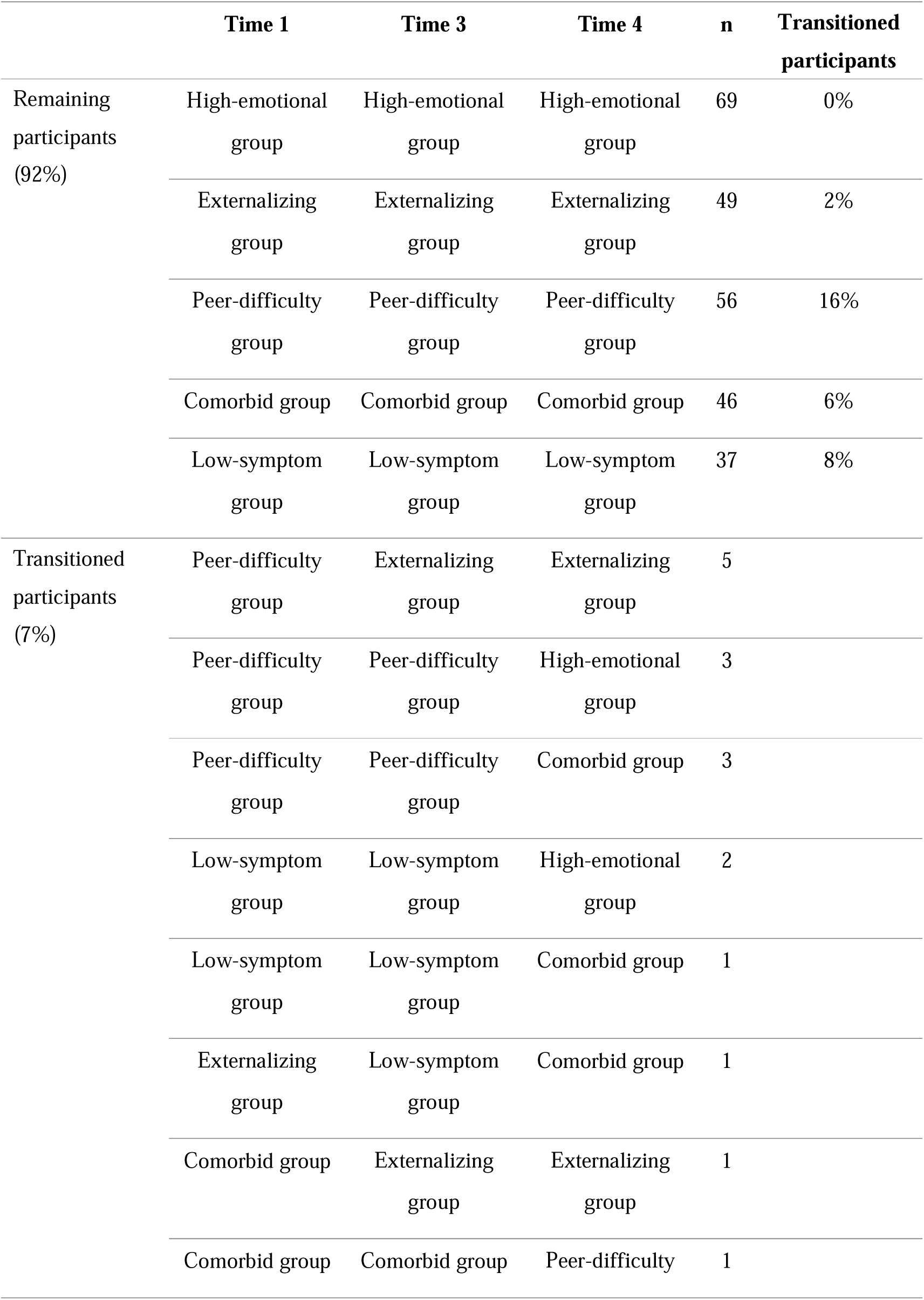
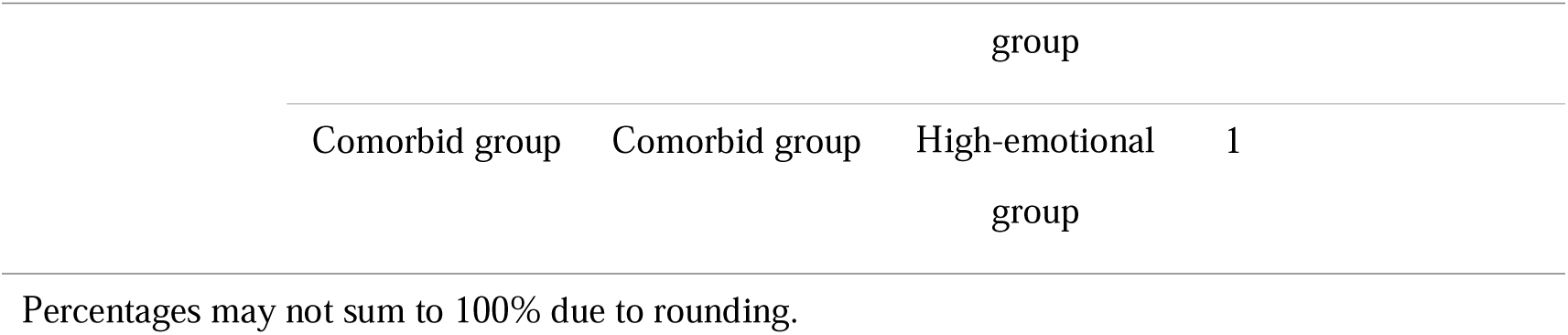
Class membership and major transition patterns.

